# The Prevalence, Etiology, And Outcome Of Anemia In Children Under Five On Admission In Three Hospitals Of Dar-Es-Salaam

**DOI:** 10.1101/2023.12.29.23300509

**Authors:** Peter Shabani Msinde

## Abstract

**Introduction:** Childhood anemia remains a persistent global challenge, emerging as the most prevalent blood disorder among children worldwide. Its enduring prevalence underscores its significance as a hematological concern of substantial public health importance, owing to its wide prevalence and potential severity. Anemia’s impact is particularly pronounced among children and expectant mothers. On a global scale, the World Health Organization (WHO) has taken significant measures to address this issue. These include implementing strategies such as iron supplementation, fortifying food sources with essential nutrients, promoting dietary diversity, and preventing conditions that contribute to anemia’s occurrence. These collective efforts aim to alleviate the burden of anemia and enhance the health and well-being of vulnerable populations.

**Objectives:** The objective of this study was to investigate the prevalence, etiologie, and clinical outcomes of anemia among children under five years of age upon admission in three district hospitals within the Dar-es-Salaam region.

**Methodology:** The study employed an observational cross-sectional design to examine the prevalence, etiologies, and outcomes of anemia in children under five who were admitted to three district hospitals in Dar-es-Salaam. Data collection took place between November 2022 and July 2023, involving children manifesting clinical signs of anemia and their caregivers. A sample size of 327 was used. Structured questionnaires were administered, and data were analyzed using SPSS software, with descriptive statistics, chi-squared tests, Fisher’s exact tests, and logistic regression employed to find associations. Ethical considerations were adhered to throughout the study.

**Results:** The study analyzed data from 327 children under five years old admitted to three district hospitals in Dar-es-Salaam to determine the prevalence, etiologies, and outcomes of anemia. The overall prevalence of anemia was found to be 45.9%, with 21.38% having mild anemia, 60% moderate anemia, and 18.62% severe anemia. Factors associated with anemia included age, referral status, family size, age of caregivers, chronic illness, deworming status, iron supplementation, active bleeding, breastfeeding, feeding adequacy, and associated comorbidities. Anemic children showed longer stays in both the ICU and wards, with an increased need for mechanical ventilators and blood transfusions.

**Conclusion and recommendation:** In conclusion, this study pinpoint the persistent burden of childhood anemia in the Dar-es-Salaam region, highlighting the need for comprehensive strategies to address its prevalence and impact. The high prevalence of anemia, particularly moderate and severe cases, suggests an important need for targeted interventions. To mitigate this issue, it is recommended that health authorities and stakeholders implement and reinforce iron supplementation programs, promote dietary diversity, enhance deworming efforts, and raise awareness about anemia’s implications. Furthermore, healthcare facilities should prioritize early detection and management of anemia in pediatric patients to reduce the associated morbidity and resource utilization.

## OPERATIONAL DEFINITIONS

### Anemia

Anemia is a medical condition characterized by a reduction in the number of red blood cells in the body, leading to a decreased ability to carry oxygen to tissues. Anemia itself is considered a symptom of an underlying health issue, rather than being a standalone diagnosis

### Etiology

Is the reasons or causes behind diseases and health problems. In medical field. It involves identifying the factors, such as genetic, environmental, or physiological, that contribute to the development and manifestation of a particular medical issue.

### Outcome

Refers to the end result or consequence of a medical treatment, procedure, or condition.

### Prevalence

is the percentage of the population that has specified diseases over period of time.

### Hemoglobin

Hemoglobin is a protein found in the blood that plays a vital role in transporting and exchanging respiratory gases, such as oxygen and carbon dioxide, between the lungs and body tissues.

## CHAPTER ONE

### 1.0 INTRODUCTION

#### 1.1 Background

Childhood anemia continue to persist worldwide. This condition is said to be the commonest blood disorder in children in the worldwide(1). According to World Health Organization a child is said to be anemic if has hemoglobin level less than 11.0 gram per deciliter(1,2). Anemia continue to be hematological disorder of major public health concern due to the fact has high prevalence as well as can present in severe form.

Among all other populations, children and expectant mothers are particularly susceptible to anemia (3). According to recent findings by World Health Organization in 2019 show that the overall prevalence of anemia 39.8% in children globally, with the African region experience the highest impact of anemia. The trend of anemia has been slowly declining internationally over the time (4). In accordance with Tanzania demographic health survey of 2015 the anemia was prevalent by 58% in children below the age of 5 years(5).

The pathophysiologic mechanism of anemia is explained by various etiological factors with three main processes involved. Loss of blood either acute or chronic, breakdown of red blood cells, and reduction of red blood cell production are reported to be main mechanism that take place in pathogenesis of this condition(6).

Tanzania has a high prevalence of nutritional anemia greatly attributed by low consumption of food with enough iron. Other causes that attribute to standstill prevalence of anemia in Tanzania include malaria and worm infestation(7).

Due to implication of anemia in quality of life of children under the age of 5, caregivers and the economic, numerous initiatives have been devised to address this issue on a global and national scale. Globally the World Health Organization has put various steps to tackle the problem which including Iron supplementation, to fortify food, ensuring dietary diversity and prevention of disorder that cause anemia(8).

At the national level government and other stakeholders have put various initiatives in fighting against anemia ranging from prenatal initiatives to postnatal initiatives. These steps includes initiation of national nutrition strategies with its component of integrated package for anemia control. Integrated package for anemia includes supplementation of iron and folate, deworming campaign, Intermittent presumptive malaria treatment, use of insecticide treated bed nets, nutritional promotion through education, environmental sanitation and hygiene(9).

#### 1.2 Statement of the Problem

Hemoglobin is an extremely significant component of red blood cells and is essential for the body’s transfer of respiratory gases. Significantly lower hemoglobin levels results in a loss in red blood cells’ ability to carry and distribute respiratory gases to and from tissues(4).

Due to its high prevalence, severe symptoms and potential connections to other nutritional issues, anemia is a public health concern(1,4). According to the World Bank, iron deficiency anemia is the most common type of anemia accounting for 50% of cases, yet it receives less attention overall, especially in middle and low income nations(4).

The most afflicted groups are children under the age of 5, and expectant mothers and the situation is made worse by the lack of access to healthcare services. This poses another huge challenge in already fragmented healthcare system. Since anemia has implications for the economy and development of the country, its effects on children’s cognitive development made it to demand more attention(10,11).

According to the World Bank, if this issue is not resolved, it will reduce the Gross Domestic Product by US Dollar 2.32 per person owing to lost productivity caused by anemia and increases the cost of living by 4.05% due to cognitive impairment (11). Given the fact that anemia effects various aspects on children life (12) this challenge requires significant effort to be invested in term of resources mobilization and prioritization.

Anemia remains to be major public health issue which require multifaceted intervention to fill the information gap as well as to improve management of these patients clinically so as to reduce mortality and morbidity in children under the age of five.

#### 1.3 Rationale

Anemia remains a standstill concern in children whereby its consequences are manifested in various aspects of life ranging from psychological aspect, physical aspect, and social aspect. Impact of anemia in cognitive has long term outcomes later on child’s life by affecting in school performance.

Due to long term and short-term implication of anemia in children it is important to evaluate how is situation of anemia in children under the age of five so as to properly intervene the situation according to vivid information generated via this research as far as etiology, prevalence and outcome of anemia in under five is concerned.

This study acted as the baseline for further studies that aimed to explore this topic. Moreover, this study generated knowledge and information would help policy makers and planners to obtain relevant information, especially concerning anemia in children under five years of age.

#### 1.4 Research Questions

1. What is the prevalence of anemia in children under age of five on admission in three district hospitals of Dar-es-salaam?
2. What are the etiologies of anemia in children under age of five on admission in three district hospitals of Dar-es-salaam?
3. What are the outcomes of anemia in children under age of five on admission in three district hospitals of Dar-es-salaam?

#### 1.5 Research Objectives

##### 1.5.1 Broad Objectives

To determine the prevalence, etiology and outcome of anemia in children under the age of five on admission in three district hospitals of Dar-es-salaam region.

##### 1.5.2 Specific Objectives

1. To determine the prevalence of anemia in children under the age of five on admission in three district hospitals of Dar-es-salaam region.
2. To determine etiologies of anemia in children under the age of five on admission in three districts hospitals of Dar-es-salaam region.
3. To determine the outcomes of anemia in children under age of five on admission in three district hospitals in Dar-es-salaam region.

#### 1.4 Conceptual framework

## CHAPTER TWO

### 2.0 LITERATURE REVIEW

#### 2.1 Prevalence of anemia in children under five

Anemia is still a major global public health issue with a high risk of affecting children and expectant mothers(13). According to the World Health Organization, 42% of children under the age of five are anemic worldwide.(13). Iron continues to be nutrient that its deficiency accounts for the majority of the nutrient related form of malnutrition worldwide (14).

According to the WHO, the Africa region has the highest prevalence of anemia in children under the age of five with 60.2% with low-quality meals being the main cause of condition’s continued prevalence. Other causes, such as being exposed to an infection, are also implicated for the issue. (15,16). High incidence has been documented in developing nations, especially in South Asia(58%) and East Africa(55%) (17).

In our nation, anemia affects 58% of the children under the age of five, according to reported data(16). According to a recent study conducted in Arusha, 84.6% of children under the age of five had anemia, with several etiologies being shown to contribute to the high prevalence(16). Anemia prevalence was reported to be 37.9% in another study in Rombo district, and it was higher in children between the ages of 6 and 23 months by 48.3% compared to those between the ages of 24 and 49 months by 28.5%(18). Different national campaigns to fight the issue have been under way to address this concern.

Additionally, a study conducted in Zanzibar revealed that anemia was prevalent with mild anemia accounting for 43.8% of cases, moderate anemia for 22.9% and severe anemia for 2.4%(19). These frightening high anemic prevalence rate need immediate action from multifaceted dimensions.

#### 2.2 Etiologies of anemia in under five children

Pregnant women and children under the age of five are disproportionately affected by anemia. Anemia can have many different causes, but the high prevalence of anemia in children under five is attributed to iron deficiency in the diet (15). Other dietary and non-nutritional factors can also contribute to anemia.(20). Infections play a role in anemia pathology particularly diseases like malaria in areas where malaria is endemic (17). Malaria was observed to significantly lower Hb levels by an average of 7.5g/dl (21). Another study show that anemia is among the predicted factor for death in children diagnosed with malaria with Odd ratio of 0.76.(22)

Also, hematological diseases like Sickle cell diseases attribute to anemia. According to 2018 study conducted in Mbeya, the prevalence of sickle cell anemia was reported to be 18% (23). Anemia was the one factor contributing to 36.9%of the hospital admissions in a systemic review study conducted at Muhimbili National Hospital (24)

Furthermore, worm infestations contribute to the pathology of anemia. A study conducted in Nigeria reveals a strong correlation between worm infestation and anemia in children under the age of five.(15,25)

#### 2.3 The hemoglobin level among under five

According to WHO standard criterion, children are deemed anemic if their Hemoglobin level is less than 11g/dl.(26)

The cut-off thresholds are as follows:10.0-10.9g/dl is mild anemia, 7.0-9.9g/dl is moderate anemia, and less than 7g/dl is severe anemia (27). In term of percentiles, anemia is defined as having hemoglobin levels below the age 5^th^ percentile (28)

The Hb level is influenced by a number of dietary and non-dietary factors, including those related to children, mothers or caregivers or even household levels (29). One study done in under five in Timor-Leste study show that mean hemoglobin levels that were higher for female(11.9g/dl) than for boys(11.7g/dl)(30).

Another study which was done in Ethiopia found that Hb level to be higher in those children aged 6-23 months old than those 24-59 months (31). Another study done in China reveals a considerable gender difference in Hb level between boys and girls in younger children, but not in older children.(32)

#### 2.4 Co-morbidities associated with anemia

Numerous illnesses have been documented to coexist with anemia and can even impair the prognosis of the patient. Stunting and other form of malnutrition can coexist with anemia. Chronic malnutrition like stunting is frequently documented in anemic children (33).

Anemia is presented also in chronic diseases which may be non-specific thus making diagnosis extremely difficult difficult.(34) (35)

Children who are anemic have been linked to condition like pneumonia(36). According to a study conducted in Bangladesh, children with pneumonia had a significant higher prevalence of anemia.(37)

In rural areas with poor sanitation, anemia is also highly common, which increases the risk of diarrhea in young children under five. These kids frequently get anemia and diarrhea.(39)

#### 2.5 Clinical outcome of children under five who with anemia

Children who anemic before the age of five frequently experience worse outcomes than non-anemic children. Clinical outcomes in pneumonic children with anemia included respiratory failures and death.(30)(37)

According to a Chinese study, anemia increases the likelihood of a child being admitted to an intensive care by 16.8% compared to non-anemic by 3.6%. Anemia also increases the likelihood of endotracheal intubation by 11.0% compared to 1.3% in non-anemic cases and it increases the likelihood of empyema by 8.6% compared to 0.6% in non-anemic cases (32)

In-hospital mortality was shown to be 12.2% in one study done in Tanzania to assess the prognosis of severe anemia following hospitalization, in contrast to that post-hospital mortality was shown to be higher by 20.2% (40)

Anemia patients were 1.55 times more likely to be hospitalized than non-anemic children with chronic kidney diseases, according to a study. According to a different study, anemic patients are increasingly being admitted to hospitals and increases need of hospital resources like blood transfusions and intensive care.(41)

These demonstrate that anemia is still a marker of poor clinical outcome for young patients who present to hospital under the age of five.

## CHAPTER THREE

### 3.0 METHODOLOGY

#### 3.1 Study design

This study was an observational cross-sectional study.

#### 3.2 Study area

This study was conducted at the Emergency Medicine departments of three selected hospitals: Muhimbili National Hospital, Temeke Regional Referral Hospital, and Amana Regional Referral Hospital. Muhimbili National Hospital, located in the Dar-es-salaam region, is a tertiary national hospital with a bed capacity of 1500, offering specialized and super specialized services. Temeke hospital serves as a regional referral hospital in the Eastern part of Dar-es-salaam, originally established in 1972 and later upgraded. Amana hospital, founded in 1954, underwent upgrades in 1982 and 1990 to become a district and then a regional referral hospital in 2010.

#### 3.3 Study duration

This study was conducted from November 2022 to July 2023.

#### 3.4 Study Population

The study population included all children under the age of five who attended the emergency department at Muhimbili National Hospital, Temeke, and Amana.

#### 3.5 Sample size calculation

The sample size was calculated using the following formula;

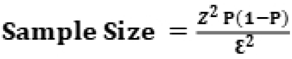

Where n is the minimum sample size required Z is standard normal deviation which is 1.96

P is assumed to be 26.2% which is proportion of anemia in under five (From the study done in Bushenyi District Western Uganda by Kikafunda et al)

E is margin of error which is 5%.

Therefore

n = 1.96^2^ x 0.262 x (1-0.262) /0.05^2^

n =297

Adjusting the non-response Assume non response rate is 10%

Therefore 10% of 297 is 29.7 which is estimated to 30

Therefore, the sample size after adjustment for the non-response was 327.

#### 3.6 Study Variables

##### 3.6.1 Independent Variables

i. Sociodemographic characteristics of the child.
ii. Sociodemographic characteristics of the caregivers/mother.
iii. Medical history of the patient.
iv. Dietary and nutritional information of the child.
v. Anemia status of the child.
vi. Clinical profile of the current illness.

##### 3.6.2 Dependent Variable

i. Clinical outcome of anemia.

#### 3.7 Inclusion Criteria AND Exclusion Criteria

##### 3.7.1 Inclusion Criteria

- Children under the age of five who have been admitted to emergency departments of hospitals during the study period.
- Those children who exhibit clinical signs of anemia, including mild or severe palmar pallor.

##### 3.7.2 Exclusion Criteria

- Children whose caregivers or mothers declined to provide consent for their participation in the study.
- Children who were confirmed to be deceased upon arrival at the hospital or during the course of treatment.

#### 3.8 Sampling

A consecutive sampling was used to enroll all participants who met the criteria until the minimum required sample size was reached. Clinical diagnosis of anemia was made based on laboratory findings whereby the Hb of less than 11g/dl was considered anemic.

#### 3.9 Data collection

Structured questionnaires were administered to gather information from the mothers or caregivers of children under the age of five who had been admitted to the emergency department. The questionnaire comprised sections that evaluated both the patients’ and the mothers’ or caregivers’ sociodemographic details. Additional sections encompassed the patient’s medical history, the child’s dietary and nutritional particulars, the child’s anemia status, the clinical presentation of the ongoing illness, and the ultimate outcome of the patient.

#### 3.10 Data analysis plan

Upon the completion of data collection, the collected data was inputted into the specialized software for data analysis, namely SPSS version 23. Subsequently, the process proceeded with data cleaning, followed by the analysis phase. In this analysis, descriptive statistics were employed to succinctly summarize the gathered data. Categorical data were presented as proportions or frequencies, while numerical variables were summarized using measures of central tendency such as mean and median. To examine the relationships between categorical variables, the chi-squared test or Fisher’s exact test was employed based on appropriateness. Furthermore, logistic regression was utilized to evaluate the connection between independent and dependent variables. The resulting data was visually presented through the use of figures, tables, and narrative descriptions to ensure a comprehensible representation. In terms of statistical significance, a P-value below 5% was considered indicative of significance for the analyzed data.

#### 3.11 Ethical consideration

Institutional review board of Muhimbili University of health and Allied Science gave ethical approval for this work in Dar es Salaam, Tanzania. Ethical approval was also secured from the Institutional Review Boards of Muhimbili National Hospital, Temeke Hospital, and Amana Hospital. Participants were assigned special codes to protect anonymity, and no personally identifiable information was collected. The study adhered to informed consent procedures, and participants were informed about the risks and benefits.

## CHAPTER FOUR

### 4.0 RESULTS

#### 4.1 Socio-demographic characteristics of participants (child and caregiver)

The study consisted 327 children with age distribution ranged from 6 to 50 months, with the majority falling within the 6-11 months and 12-23 months age groups. The gender distribution of children was approximately balanced, with 53.2% being male and 46.8% female. Urban residence was more prevalent at 56.9% compared to rural at 43.1%. A majority of children (85.3%) had not yet started school. Self-referral was the most common referral status (30.6%), and uninsured (50.2%) were slightly higher than insured (49.8%). Most caregivers were between 25-34 years (53.5%) and married (94.5%). Most of mothers/caregivers had primary education (38.2%), were self-employed (17.1%), and majority lived in nuclear households (85.9%). More details in table 1.

**Table 1:**
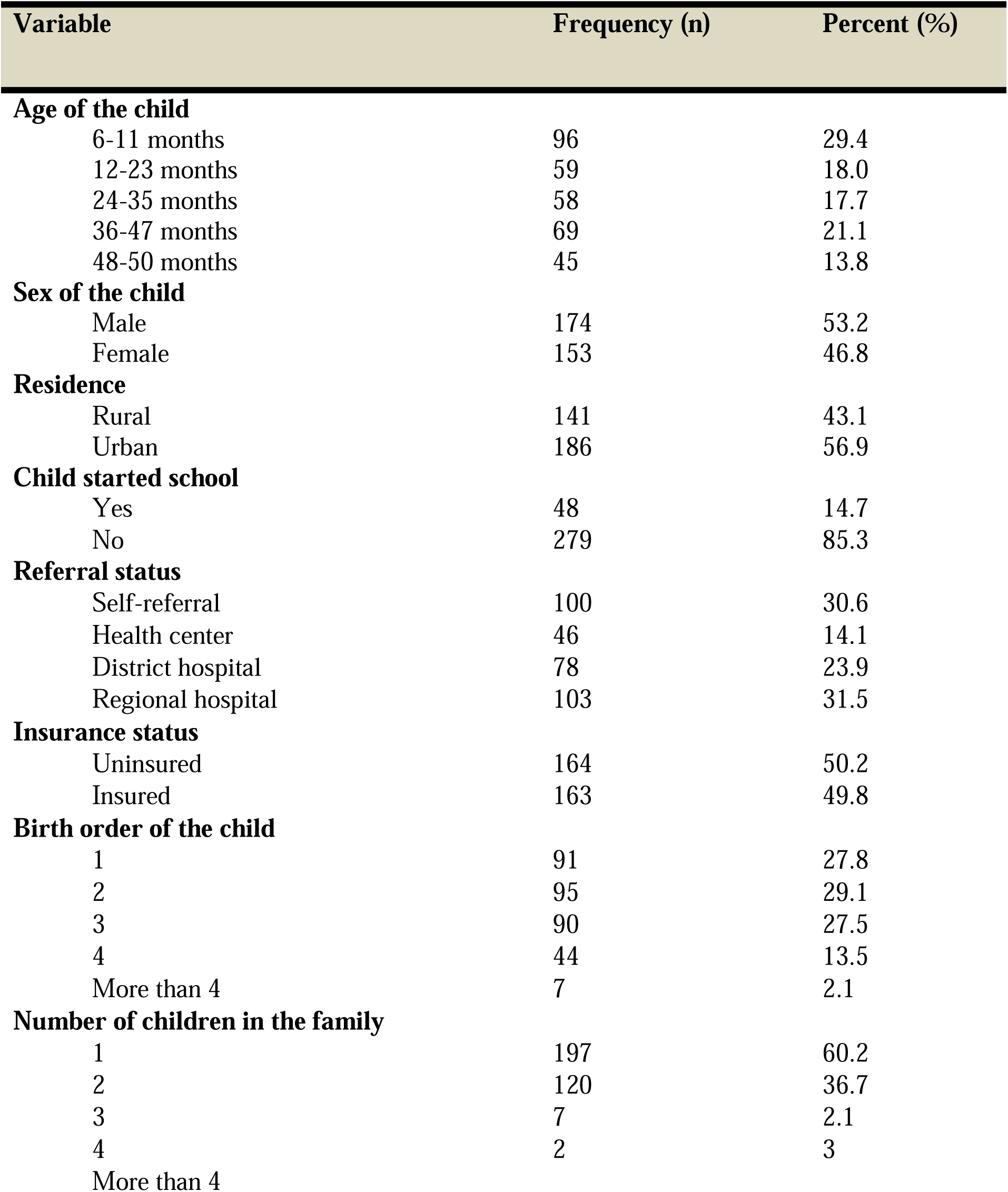
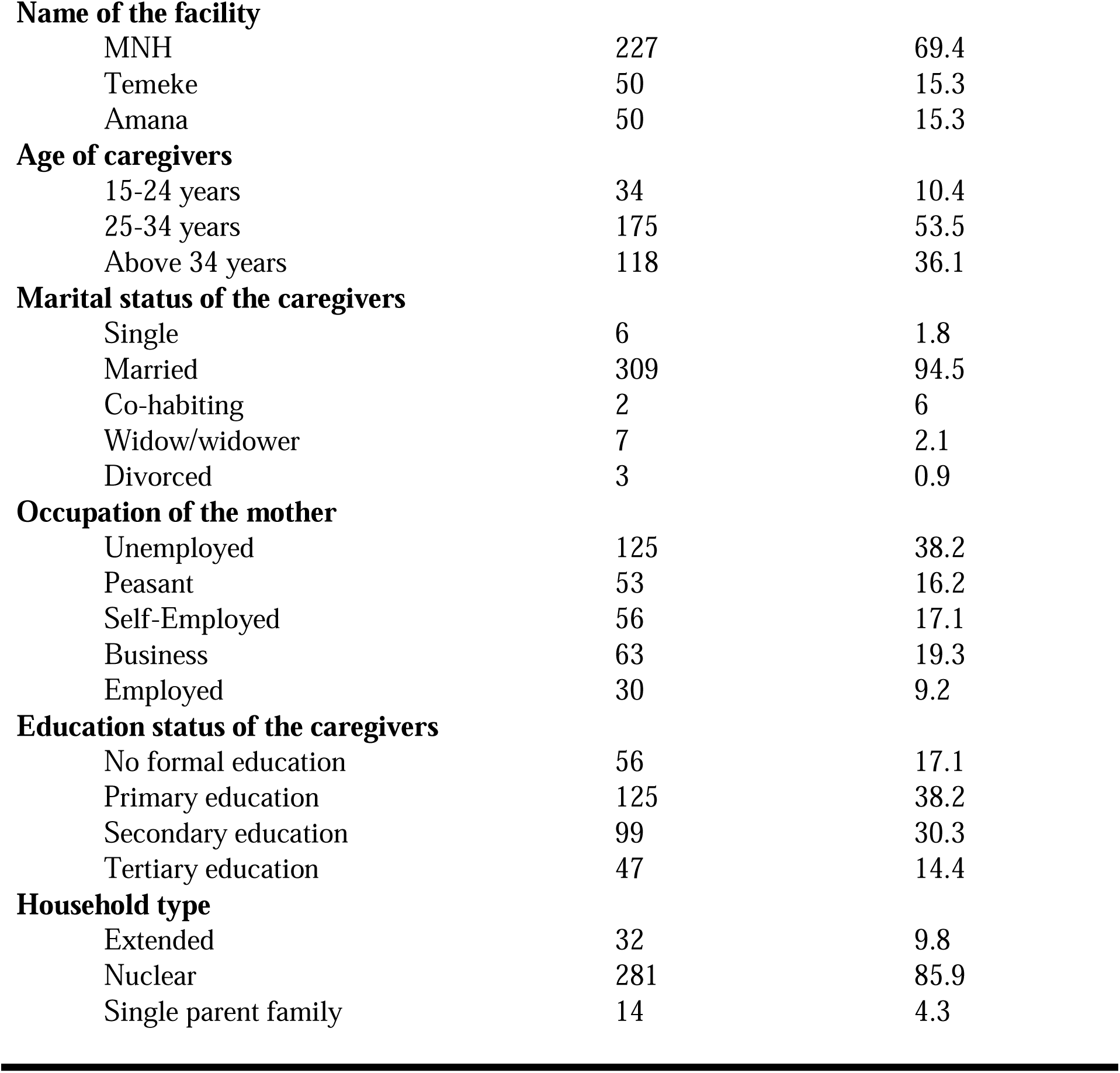
Socio-demographic characteristics of participants (n=327)

#### 4.2 Prevalence of anemia in under five children

The study aimed to assess the prevalence of anemia among children under the age of five in three referral hospitals. Anemia was determined based on the hemoglobin levels measured through laboratory tests conducted in both the Emergency department or the hospital wards. Hemoglobin levels below 11g/dl were used as the cut-point for diagnosing anemia. The study revealed that the overall prevalence of anemia among the children was found to be 45.9%. A more detailed in Figure 1 below.

**Figure 1:**
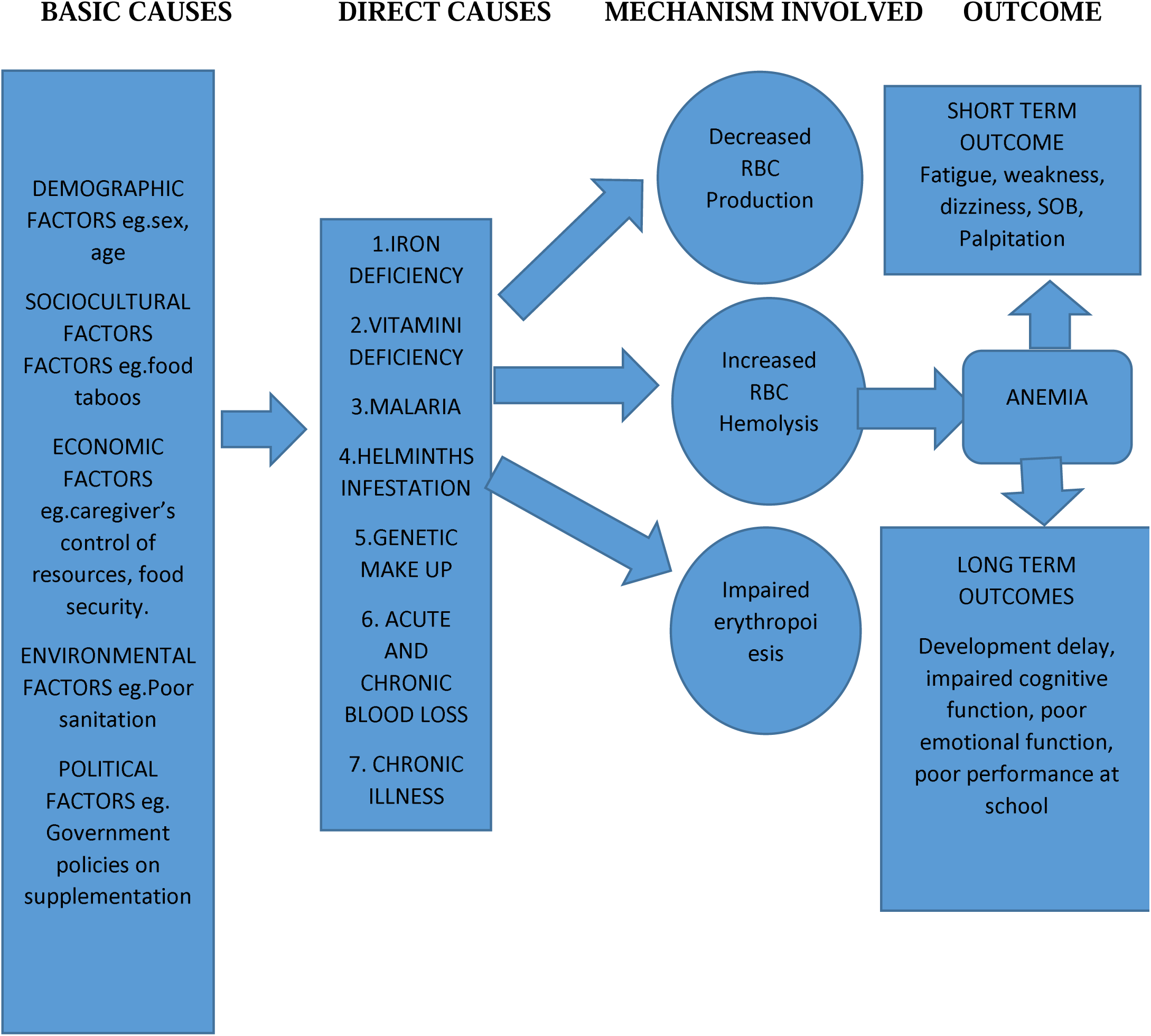
Conceptual framework of anemia etiology and outcomes.

The study also assessed the severity of anemia which indicated that 21.38% had mild anemia (10.0-10.9 g/dl), 60% with moderate anemia (7.0-9.9 g/dl), and 18.62% with severe anemia (<7.0 g/dl). More details refer to figure 2 below

**Figure 2.**
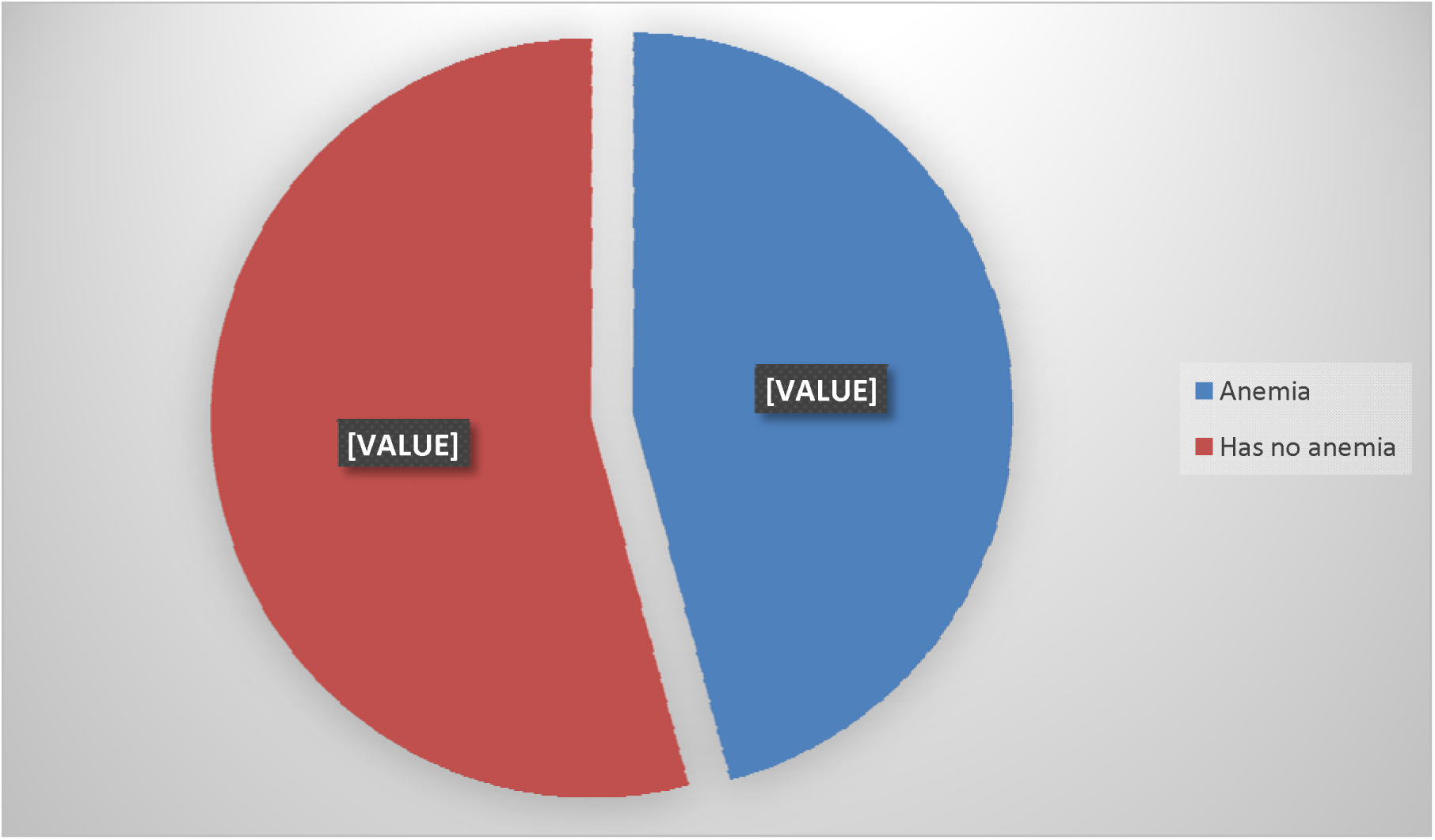
Overall prevalence of anemia in under-five children.

**Figure 3:**
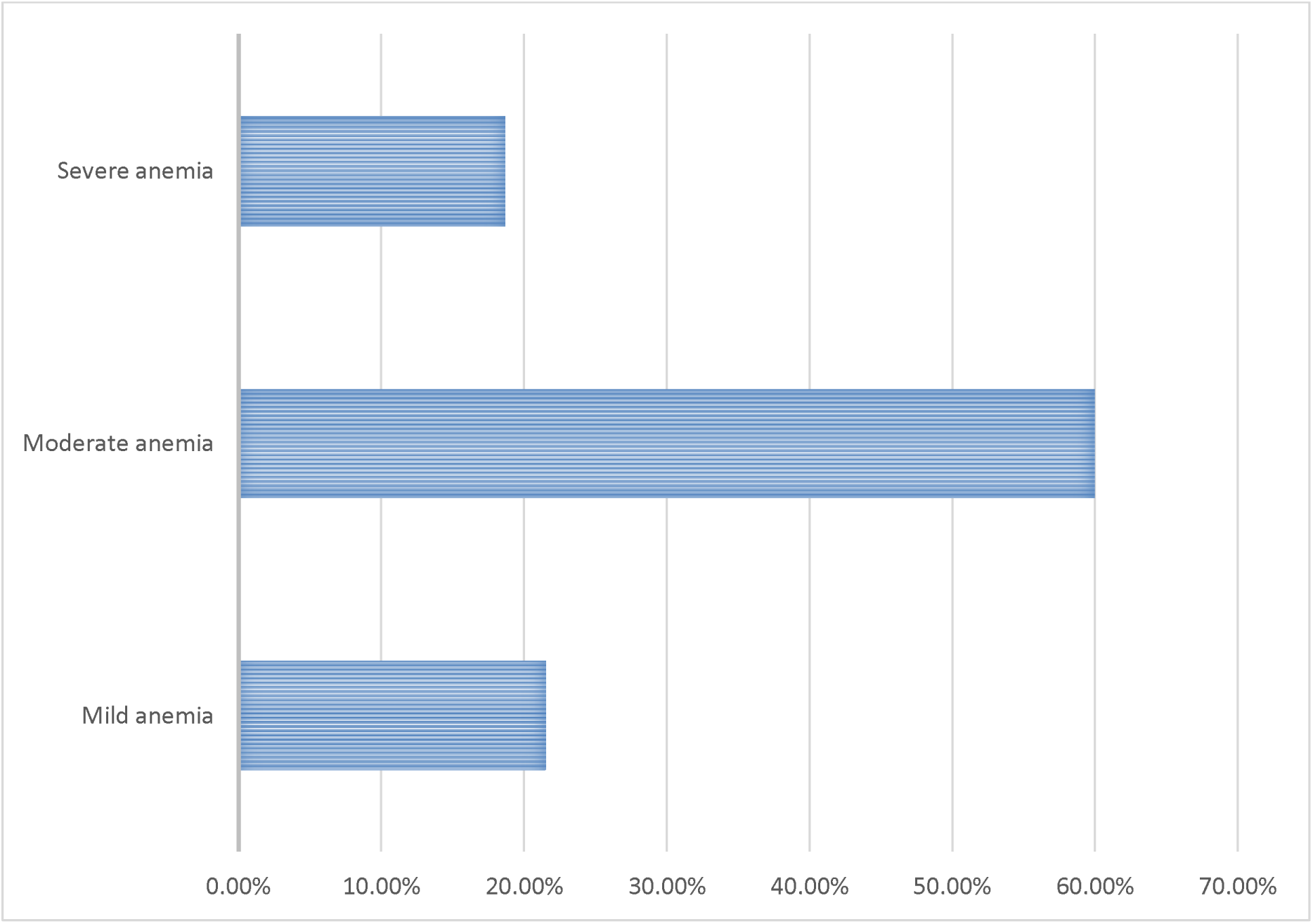
Severity of anemia.

#### 4.3 Etiologies and associated factors of anemia

The analysis revealed that various factors were associated with anemia as the following, Children aged 36-47 months showed a higher prevalence of anemia (31.9%) compared to those aged 6-11 months (49.0%) or 12-23 months (47.5%). Similarly, children referred from health centers demonstrated a lower prevalence of anemia (21.7%) compared to those referred through other facilities. Childbirth order, family size, age of caregivers, marital status, chronic illness, deworming status, iron supplementation, active bleeding, breastfeeding, feeding adequacy, and associated comorbidities also showed significant associations with anemia prevalence with P-value less than 0.05. Anemia appears to be more in hematological diseases (64.2%) and infectious diseases (52.6%), suggesting potential relation between anemia and these two conditions. Also, neurological diseases showed a significant difference in prevalence between anemic and non-anemic cases (42.2% vs. 57.8%). More information in table 3 below.

**Table 2:** Associated comorbidities and signs and symptoms of anemia.

| Variable | Frequency (n) | Percent (%) |
| --- | --- | --- |
| **Associated comorbidities/reason for admission** |  |  |
| Infectious | 76 | 23.2 |
| Hematological diseases | 53 | 16.2 |
| Malignancy | 9 | 2.8 |
| Neurological diseases | 64 | 19.6 |
| Congenital diseases | 28 | 8.6 |
| Respiratory diseases | 25 | 7.6 |
| Malnutrition | 51 | 15.6 |
| Renal diseases | 19 | 5.8 |
| Others | 2 | 6 |
| **Symptoms of anemia** |  |  |
| Lethargy | 58 | 17.7 |
| Irritability | 21 | 6.4 |
| Poor feeding | 30 | 9.2 |
| Yellowish coloration | 5 | 1.5 |
| Failure to thrive | 21 | 6.4 |
| Poor attention span | 3 | 0.9 |
| Poor sleeping associated with excessive crying | 3 | 0.9 |
| None | 186 | 56.9 |
| **Signs of anemia** |  |  |
| Pallor | 108 | 33.0 |
| Nails become thins, brittle, flat. | 4 | 1.2 |
| Atrophy of the tongue | 2 | 0.6 |
| Tachycardia | 17 | 5.2 |
| Tachypnea | 4 | 1.2 |
| Splenomegaly | 4 | 1.2 |
| None | 188 | 57.5 |

**Table 3:** Etiology and factors associated with anemia in under five children.

| Variable | Anemia<br>N (%) | No anemia<br>N (%) | P value |
| --- | --- | --- | --- |
| **Age of the child** |  |  |  |
| 6-11 months | 47(49.0) | 49(51.0) | 0.103 |
| 12-23 months | 28(47.5) | 31(52.5) |  |
| 24-35 months | 28(48.3) | 30(51.7) |  |
| 36-47 months | 22(31.9) | 47(68.1) |  |
| 48-50 months | 25(55.6) | 20(44.4) |  |
| **Sex of the child** |  |  |  |
| Male | 83(47.7) | 91(52.3) | 0.479 |
| Female | 67(43.8) | 86(56.2) |  |
| **Residence** |  |  |  |
| Urban | 60(42.6) | 81(57.4) | 0.294 |
| Rural | 90(48.4) | 96(61.6) |  |
| **Child started school** |  |  |  |
| Yes | 28(58.3) | 20(41.7) | 0.061 |
| No | 122(43.7) | 157(56.3) |  |
| **Referral status** |  |  |  |
| Self-referral | 40(40.0) | 60(60.0) | <0.001 |
| Health center | 10(21.7) | 36(78.3) |  |
| District hospital | 33(42.3) | 45(57.7) |  |
| Regional hospital | 67(65.0) | 36(35.0) |  |
| **Insurance status** |  |  |  |
| Insured | 77(47.0) | 87(53.0) | 0.694 |
| Uninsured | 73(44.8) | 90(55.2) |  |
| **Birth order of the child** |  |  |  |
| 1 | 40(44.0) | 51(56.0) | 0.264 |
| 2 | 45(48.4) | 49(51.6) |  |
| 3 | 46(51.1) | 44(48.9) |  |
| 4 | 14(31.8) | 30(68.2) |  |
| More than 4 |  |  |  |
| **Number of children in the family** |  |  |  |
| 1 | 75(38.1) | 122(61.9) | 0.009 |
| 2 | 68(56.7) | 52(43.3) |  |
| 3 | 5(71.4) | 2(28.6) |  |
| 4 | 1(50.0) | 1(50.0) |  |
| More than 4 | 1(100.0) | 0(0.0) |  |
| **Age of caregivers** |  |  |  |
| 15-24 years | 9(26.5) | 25(73.5) | 0.001 |
| 25-34 years | 98(56.0) | 77(44.0) |  |
| Above 34 years | 43(36.40) | 75(63.6) |  |
| **Marital status of the caregivers** |  |  |  |
| Single | 2(33.3) | 4(66.7) |  |
| Married | 137(44.3) | 172(55.7) |  |
| Co-habiting | 2(100.0) | 0(0.0) | 0.026 |
| Widow/widower | 6(85.7) | 1(14.3) |  |
| Divorced | 3(100.0) | 0(0.0) |  |
| **Occupation of caregivers** |  |  |  |
| Unemployed | 66(52.8) | 59(47.2) |  |
| Peasant | 23(43.4) | 30(56.6) |  |
| Self-Employed | 23(41.1) | 33(58.9) | 0.403 |
| Business | 26(41.3) | 37(58.7) |  |
| Employed | 12(40.00) | 18(60.0) |  |
| **Education status of the caregivers** |  |  |  |
| No formal education | 31(55.4) | 25(44.6) |  |
| Primary education | 56(44.8) | 69(55.2) | 0.469 |
| Secondary education | 43(43.4) | 56(56.6) |  |
| Tertiary education | 20(42.6) | 27(57.4) |  |
| **Household type** |  |  |  |
| Extended | 19(59.4) | 13(40.6) |  |
| Nuclear | 121(43.1) | 160(56.9) | 0.031 |
| Single parent family | 10(71.4) | 4(28.6) |  |
| **Diagnosis for admission/Associated comorbidities** |  |  |  |
| Infectious | 40(52.6) | 36(47.4) |  |
| Hematological diseases | 34(64.2) | 19(35.8) |  |
| Malignancy | 3(33.3) | 6(66.7) |  |
| Neurological diseases | 27(42.2) | 37(57.8) | 0.010 |
| Congenital diseases | 8(28.6) | 20(71.4) |  |
| Respiratory diseases | 5(20.0) | 20(80.0) |  |
| Malnutrition | 22(43.1) | 29(56.9) |  |
| Renal diseases | 10(52.6) | 9(47.4) |  |
| Others | 1(50.0) | 1(50.0) |  |
| **Deworming status** |  |  |  |
| Yes | 74(37.8) | 122(62.2) | 0.001 |
| No | 76(58.0) | 55(42.0) |  |
| **Iron supplementation** |  |  |  |
| Yes | 13(86.7) | 2(13.3) | 0.001 |
| No | 137(43.9) | 175(56.1) |  |
| **Active bleeding** |  |  |  |
| Yes | 55(30.7) | 124(69.3) | 0.001 |
| No | 95(64.2) | 53(35.8) |  |
| **Family history of blood disorder** |  |  |  |
| Yes | 21(40.4) | 31(59.6) | 0.387 |
| No | 129(46.9) | 146(53.1) |  |
| **Adequate breastfeeding** |  |  |  |
| Yes | 107(45.5) | 128(54.5) |  |
| No | 26(61.9) | 16(38.1) | 0.027 |
| Not applicable | 17(34.0) | 33(66.0) |  |
| **Adequate feeding** |  |  |  |
| Yes | 58(44.3) | 73(55.7) | 0.004 |
| No | 99(100.0) | 0(0) |  |

#### 4.4 Clinical outcome of anemia

Regarding the outcome of illness, no significant differences were observed between anemic and non-anemic children in terms of death, admission to the Pediatric Intensive Care Unit (PICU), readmission, or discharge home improved. However, more anemic children were admitted to the ward compared to non-anemic children. The duration of stay in both the ICU and wards was significantly different between anemic and non-anemic children, with a larger proportion of anemic children staying for more than 14 days. Additionally, anemia was associated with a greater need for mechanical ventilators and blood transfusions. More details in table 4.

**Table 4:** Clinical outcome of anemia (n=327)

|  | Anemic<br>N (%) | No<br>anemia<br>N (%) | P-value |
| --- | --- | --- | --- |
| Outcome variables |  |  |  |
| Outcome of illness |  |  |  |
| Death | 19(12.7) | 23(13.0) | 0.289 |
| Admitted to PICU | 12(8.0) | 18(10.2) |  |
| Readmission | 19(12.7) | 20(11.3) |  |
| Discharge home improved | 0(0.0) | 5(2.8) |  |
| Admitted in the ward | 106(66.7) | 111(62.7) |  |
| Number of days in ICU |  |  |  |
| Less than 14 | 17(11.3) | 2(1.1) | 0.001 |
| More than 14 | 3(2.0) | 0(0.0) |  |
| Not applicable | 130(86.7) | 175(98.9) |  |
| Numbers of days in the wards |  |  |  |
| Less than 14 | 90(60.00) | 112(63.30) | 0.605 |
| More than 14 | 41(27.3) | 40(22.6) |  |
| Not applicable | 19(12.70) | 25(14.1) |  |
| Total hospitalization days |  |  |  |
| Less than 14 | 87(58.0) | 108(61.0) | 0.551 |
| More than 14 | 45(30.0) | 44(24.9) |  |
| Not applicable | 18(12.0) | 25(14.1) |  |
| Need for mechanical ventilators |  |  |  |
| Yes | 21(14.0) | 2(1.1) | 0.001 |
| No | 2(1.1) | 175(98.9) |  |
| **Need for blood transfusion** |  |  |  |
| Yes | 28(19.7) | 0(0.0) | 0.001 |
| No | 122(81.3) | 177(100.0) |  |

## CHAPTER FIVE

### 5.0 DISCUSSION

#### 5.1 Prevalence of anemia in children under the age of five on admission in three district hospitals of Dar-es-salaam region

This rstudy investigated the prevalence, causes, and outcomes of anemia among children under five years old who were admitted to three hospitals in Dar es Salaam. The overall prevalence of anemia was recorded at 45.9%. This finding closely resembled the anemia prevalence reported for the Amhara Region in the 2016 Ethiopian DHS (42%) (37) and a study conducted in Jimma, Ethiopia (46%) (38). Nevertheless, the outcome of this current study was lower than the rates observed in South-East Nigeria (49.2%)(39) Hohoe municipality and Volta Regional Hospital in Ghana (47.5% & 55.0%) (40) (41) and Limpopo Province, South Africa (75.0%) (42)

This divergence in prevalence could be attributed to variations in the design of the studies, sampling methods, and sample sizes. Additionally, differences might stem from the geographical locations of the participants or disparities in socio-demographic characteristics and the socioeconomic status of parents within those regions.

Concerning the severity of anemia, the majority of anemic children in this study exhibited moderate anemia (60%), followed by mild anemia (21.38%), and severe anemia (18.62%). This finding concurs with research conducted in Nepal (Mathema S and Shrestha A,) as well as in Hohoe municipality, Ghana (40) and in India (43) Furthermore, our findings align with the 2016 EDHS report (37) and the Volta Regional Hospital study in Ghana (41) which both indicated a notable prevalence of moderate anemia.

The medical experts in the study successfully employed clinical indicators to recognize the majority of cases involving severe anemia as well as the combined category of “moderate or mild” anemia. Additionally, they generally managed to distinguish between these levels of anemia and instances where no anemia was present. In earlier research (44)(45) overlapping degrees of paleness were utilized to identify severe and moderate anemia. In contrast, our study employed pronounced pallor to classify severe anemia and some pallor to categorize “moderate or mild” anemia. This differentiation is crucial for the accurate detection and appropriate management of both levels of anemia.

The use of severe pallor alone displayed relatively modest to reasonable sensitivity and high specificity for the identification of severe anemia. The inclusion of the symptom of lethargy alongside an “or” condition augmented the identification of severe anemia. However, this modification negatively impacted the specificity and positive predictive value associated with severe pallor’s indication of severe anemia, as well as the diagnosis of “moderate or mild” anemia. Within this study, lethargy retained a reasonable predictive value for severe anemia diagnosis while minimizing any detrimental influence on the diagnosis of “moderate or mild” anemia. This suggests that adding lethargy to the Integrated Management of Childhood Illness (IMCI) guidelines for anemia diagnosis could enhance the identification of severe anemia cases requiring referral to a hospital.

#### 5.2 Etiologies of anemia in children under the age of five on admission in three districts hospitals of Dar-es-salaam region

The origins of anemia often stem from a complex mix of factors that interact in a multifaceted manner. To begin with, the significance of each factors such as hookworm infestations or malaria infections varies across different contexts. Anemia can manifest as a chronic condition, like when it arises from iron deficiency, human immunodeficiency virus (HIV) infections, or intestinal worm infestations. Conversely, it might appear as an acute episode due to factors like a sickle-cell crisis or Plasmodium falciparum infection. Furthermore, instances of chronic anemia can be acutely intensified. This complexity is further compounded by the fact that childhood anemia can result not just from events during childhood but also from maternal iron deficiency and anemia. These maternal conditions are linked to impaired fetal development, as well as the birth of iron-deficient and anemic babies.

Additionally, socioeconomic status can influence anemia risk by impacting nutritional status, family size, birth intervals, and by exacerbating issues of affordability and accessibility of preventive and curative measures, as highlighted (46)

Within this study, it was discovered that deworming status and iron supplementation significantly affected the likelihood of acquiring anemia. Children who received deworming treatment were less susceptible to anemia compared to those who were not dewormed. This finding underscores the role of worm infestations as a contributing factor to anemia in children.

The research indicated a higher prevalence of anemia among children under 3 years old, with the prevalence decreasing as children’s age increased. This finding aligns with similar studies conducted in Ethiopia (37)(47)(48). As well as studies in other developing countries(40)(49)(50) This pattern might be attributed to the elevated iron demands during rapid growth and erythropoiesis, insufficient iron availability in diets, and limited maternal iron reserves during pregnancy, as discussed(51).

Comparable to findings in South-East Nigeria(39) and the Volta Regional Hospital of Ghana (41). In the current investigation, no significant link was observed between gender and anemia. Conversely, alternative studies have indicated a greater occurrence of anemia among boys compared to girls(52) (47). Conversely, some studies have reported a higher prevalence of anemia in girls compared to boys,(42) These conflicting outcomes could potentially be attributed to societal norms influencing varying consumption of iron-rich foods between genders. Nonetheless, further research is imperative to gain a more comprehensive comprehension of this intricate matter

Furthermore, within the current study, the prevalence of anemia was notably higher in patients originating from rural areas, at 48.4%, in comparison to those dwelling in urban settings, at 42.6%. This discrepancy is exemplified by the higher anemia prevalence observed in the rural Bulange sub-county, reaching 79.0%, in contrast to Namutumba and Magada sub-counties. This divergence might be attributed to greater levels of illiteracy and reduced access to healthcare services, including health education (53)

The research also indicated that family size played a significant role in predicting anemia among children under five in Africa. Children hailing from households with one child or fewer than two were 38% less prone to anemia in comparison to their counterparts. This could be attributed to the correlation between larger family sizes and food insecurity. Smaller families are more likely to afford an adequate and diversified diet, rich in iron, which might explain this pattern. This finding aligns with the study done by Tadese et al,2022 (54) have previously associated high maternal parity with anemia, as increased child numbers often hinder the ability to adequately nourish them.

The age of caregivers also emerged as a statistically significant factor in the development of childhood anemia. This echoes the results of a study by Finlay et al., where young mothers with one to three children were common, and two-thirds of the children from these young mothers had anemia. Young mothers typically encounter difficulties in childcare due to limited resources and experience, potentially leading to poorer child health outcome (55).

Furthermore, the study unveiled a higher prevalence of anemia among children introduced to complementary foods before six months of age, as opposed to those exclusively breastfed for the recommended six months. Early introduction of complementary foods before six months reduces iron bioavailability by up to 80%. The introduction of foods like cow’s milk interferes with iron absorption from breast milk due to excessive protein and mineral content, notably calcium. Digestive enzymes are also insufficient at this age. Furthermore, this early introduction increases the risk of microbial contamination and subsequent diarrheal diseases, which can lead to malabsorption, (37) (56). Interestingly, this discovery contrasts with research from Sri Lanka, where children exclusively breastfed for six months or more were more likely to be anemic than those breastfed exclusively for less than six months, as observed (57)

The primary approach employed to elevate hemoglobin levels and diminish anemia is iron supplementation. Given the substantial occurrence of both iron deficiency (ID) and anemia among children in developing nations, a suggestion has been made to universally supplement children starting at six months of age. (58)(59) However, concerns have arisen regarding the safety of implementing universal iron supplementation in regions where malaria is prevalent. Interestingly, our study contradicts this notion, as it revealed that children who underwent iron supplementation exhibited higher likelihoods of developing anemia. This disparity was statistically significant in our findings.

#### 5.3 Outcomes of anemia in children under age of five on admission in three district hospitals in Dar-es-salaam region

Furthermore, this study has identified a range of outcomes that become evident in children afflicted with anemia. These outcomes encompass variables like mortality, admission to the Pediatric Intensive Care Unit (PICU), frequency of readmissions, improvement following discharge, placement within the hospital ward, duration of stay in the ICU, length of time spent in general wards, total days of hospitalization, necessity for mechanical ventilation, and the need for blood transfusion.

Within this research, the proportion of anemic children who experienced mortality was 12.7%, while for their non-anemic counterparts, this figure stood at 13.0%. The divergence in death rates between the two groups is minimal, and the associated p-value (0.289) underscores the absence of statistical significance in this difference. However, the categorization of hemoglobin (HB) levels delineates the variations within the realm of anemia. This discovery is aligned with multiple studies, emphasizing that anemia can contribute to mortality directly, particularly in severe cases, or indirectly, particularly when anemia is mild or moderate.(60) supports the notion that a significant proportion of anemia-related deaths stem from mild and moderate anemia. Given the demonstrated graded risk relationship, even a slight enhancement in hemoglobin concentration could potentially lead to a reduction in mortality rates among infants and young children.

Numerous studies have concentrated on children with extended stays in the PICU, as this subgroup is most likely to benefit from interventions tailored to PICU care such as blood conservation protocols or erythropoietin therapy. Despite representing about 20% of PICU admissions, children with PICU stays exceeding 48 hours contribute disproportionately to PICU resource utilization.(61) our findings echo this pattern. Anemic children are admitted to the PICU at a rate of 8.0%, while non-anemic children are admitted at a rate of 10.2%. Once again, the dissimilarity lacks statistical significance (p-value = 0.289). It’s worth noting that anemic children have a higher percentage (11.3%) of ICU stays lasting less than 14 days compared to non-anemic children (1.1%). This divergence is statistically significant (p-value = 0.001).

The reasons behind this could involve anemia-related complications necessitating longer ICU durations. Interestingly, the not applicable category encompasses most children, both anemic and non-anemic, implying that ICU care might not have been essential for a considerable number of cases.

Within this investigation, both anemic and non-anemic children exhibit comparable readmission rates, standing at 12.7% and 11.3% respectively. The calculated p-value signifies the absence of a noteworthy association (p-value = 0.289). However, it’s noteworthy that no anemic children demonstrated improved status upon discharge, while a modest 2.8% of non-anemic children did. Due to the limited percentage, deriving definitive conclusions from this data is challenging.

In contrast, 66.7% of anemic children and 62.7% of non-anemic children were admitted to the hospital ward. The discrepancy lacks statistical significance (p-value = 0.289). The distribution of hospitalization days parallels between both groups, with no statistically significant disparities (p-value = 0.551). This implies that anemia may not exert a significant influence on the duration of hospital stays.

A notable 14.0% of anemic children necessitated mechanical ventilators, in contrast to 1.1% of non-anemic children (p-value = 0.001). Anemia can contribute to respiratory distress, requiring ventilator assistance. Comparable percentages of anemic and non-anemic children spent less than 14 days in the wards (60.0% and 63.3% respectively), with no significant distinction (p-value = 0.605). Similarly, for extended ward stays, the variation remains slight, with anemic children at 27.3% and non-anemic children at 22.6%.

The current study highlighted the need for blood transfusions, as a substantial proportion of anemic children (19.7%) necessitated blood transfusions, in contrast to none of the non-anemic children. This association holds statistical significance (p-value = 0.001), underscoring that anemia significantly elevates the likelihood of requiring blood transfusions.(62) where 49% of children with anemia received one or more red blood cell transfusions during their PICU stay, and 6% received transfusions post-PICU discharge.

## CHAPTER SIX

### 6.0 STUDY LIMITATION, MITIGATION, CONCLUSION AND RECOMMENDATION

#### 6.1 Study Limitation

1. **Limited Geographic Area:** The study was conducted in three referral hospitals within a same region (Dar es Salaam). This limited geographical scope might affect the generalizability of the findings to a broader population.
2. **Sampling and Selection Bias:** The participant pool might not represent the entire population of children under five, as the study relies on data collected from hospitals. Children who didn’t seek medical attention or were treated outside these hospitals are not accounted for, introducing a potential sampling bias. Also, other children were treated in outpatients’ clinics were not part of this study.
3. **Symptom and Sign Variability:** The symptoms and signs of anemia might be subjective and interpreted differently by healthcare providers, leading to variability in diagnosis. For example, pallor might perceive as severe pallor in one healthcare provider but not in another person.
4. **Nature of the study design:** It does not reveal causal links between independent variables and anemia. Due to constraint of resource, we were unable to measure serum ferritin concentration, soluble transferrin receptor concentration, folate levels, vitamin B12 levels, thalassemia, and G6PD deficiency; which could have helped in finding the causes of anemia.

### 6.2 Mitigation

**1. Limited Geographic Area:**

**- Mitigation:** While the study was limited to a specific geographic region, Further studies with wider scope are encouraged to be done in the further.

**2. Sampling and Selection Bias:**

**- Mitigation:** To improve representation, future studies can consider incorporating data from multiple sources, such as community health centers and outpatient clinics.

**3. Symptom and Sign Variability:**

**- Mitigation:** Clear guidelines for diagnosing anemia-related symptoms and signs, such as pallor, should be established. This could involve using objective measurement tools whenever possible to minimize subjectivity.

**4. Cross-sectional nature of the study design:**

**- Mitigation:** Further community-based studies should be conducted to have findings more representing the whole population.

#### 6.3 Conclusion

This study highlighted on the significant issue of anemia among children under five years old in the Dar-es-Salaam region. The findings revealed a notable prevalence of anemia, with an overall prevalence rate of 45.9%. This prevalence closely resembled rates reported in similar regions and countries, indicating a substantial burden of anemia affecting these young children. The study identified that moderate anemia was the most common severity level among anemic children, emphasizing the need for early detection and intervention to prevent progression to severe anemia.

The etiological factors contributing to anemia were multifaceted, involving a combination of demographic, socio-economic, and healthcare-related variables. Factors such as age, referral status, deworming status, and iron supplementation showed significant associations with anemia prevalence, highlighting potential areas for targeted interventions. Notably, younger age groups, lack of deworming treatment, and absence of iron supplementation were linked to higher anemia rates. These findings underscore the importance of comprehensive healthcare programs that address these factors to reduce anemia prevalence.

Clinical outcomes indicated that anemia had implications beyond its prevalence. Anemic children exhibited differences in hospitalization duration, need for mechanical ventilators, and blood transfusions. These findings suggest that anemia not only affects children’s health but also places an additional burden on healthcare resources. Future interventions should consider these outcomes to develop strategies for more effective anemia management and treatment.

#### 6.4 Recommendation

##### 1. Early Detection and Intervention of anemia

Timely interventions, such as nutritional supplementation and deworming treatments, can significantly contribute to preventing the progression of anemia from one form of severity to another and its associated health complications.

##### 2. Various health promotion initiatives

Given the multifaceted nature of anemia’s contributing factors, healthcare programs should adopt various health promotion programs that should address not only the medical aspects but also socioeconomic factors that contribute to anemia. Ensuring access to quality healthcare services, promoting awareness about proper nutrition, and advocating for regular deworming and iron supplementation are vital components of such initiatives.

##### 3. Targeted Interventions like mass drug administration

The identification of specific factors like age, deworming status, and iron supplementation as correlates of anemia prevalence suggests opportunities for targeted interventions. Tailored efforts that focus on vulnerable age groups, prioritize deworming treatments, and encourage iron supplementation can effectively address the root causes of anemia in the studied population.

## Data Availability

All data produced in the present study are available upon reasonable request to the authors

## LIST OF ABBREVIATION

EMD: Emergency Department
Hb: Hemoglobin
IVD: Immunization and Vaccine Program
MNH: Muhimbili National Hospital
MUHAS: Muhimbili University of Health and Allied Science
PICU: Pediatric Intensive Care Unit
P-value: Probability value
RBC: Red Blood Cell
SPSS: Statistical Package for Social Science
US: United States
WHO: World Health Organization

